# Assessment of concordance between related systematic reviews and between related guideline recommendations: protocol for a methodological survey

**DOI:** 10.1101/2022.07.11.22277498

**Authors:** Arnav Agarwal, Loai Albarqouni, Nour Badran, Nina Brax, Pooja Gandhi, Tiago Pereira, Abigail Roberts, Ola El Zein, Elie Akl

## Abstract

Independent systematic reviewers may arrive at different conclusions when analyzing evidence addressing the same clinical questions. Similarly, independent expert panels may arrive at different recommendations addressing the same clinical topics. When faced with a multiplicity of reviews or guidelines on a given topic, users are likely to benefit from a structured approach to evaluate concordance, and to explain discordant findings and recommendations. This protocol proposes a methodological survey to evaluate the prevalence of concordance between reviews addressing similar clinical questions, and between clinical practice guidelines addressing similar topics; and to identify methodological frameworks for the evaluation of concordance between related reviews and between related guidelines.

## Introduction

Independent systematic reviewers may arrive at different conclusions when analyzing the same body of evidence (1). Similarly, independent expert panels may arrive at different recommendations when interpreting evidence addressing the same clinical questions. Such discordance may be driven by different interpretation of relative benefits and harms, and differing degrees of certainty associated with an overall body of evidence. Discordance in recommendations may also be driven by different judgments of contextual factors such as values and preferences, resource availability, equity and feasibility.

Investigators have previously evaluated the degree of concordance between the findings of systematic reviews addressing similar clinical questions (2). Similarly, the degree of concordance between guideline recommendations on similar health topics, and possible reasons for discordant recommendations, have been explored (3, 4). These evaluations have used different methodological approaches. Several frameworks for evaluating discordance in systematic reviews and in guidelines have also been developed (5-7).

When faced with a multiplicity of reviews or guidelines on a given clinical topic, users of systematic reviewers and guidelines are likely to benefit from a structured approach to evaluate concordance, and to explain discordant findings and recommendations.

### Objectives and aim

Specific to systematic reviews, our objectives are:

1. To evaluate the prevalence of concordance between reviews addressing similar clinical questions; and
2. To identify methodological frameworks for the evaluation of concordance, and explanation of any discordance, between reviews addressing similar clinical questions.

Specific to clinical practice guidelines, our objectives are:

1. To evaluate the prevalence of concordance between guidelines addressing similar topics; and
2. To identify methodological frameworks for the evaluation of concordance, and explanation of any discordance, between guidelines addressing similar health topics.

## Methods

Our study design consists of a methodological survey addressing the aforementioned objectives.

### Eligibility criteria

We will include empirical studies (i.e. direct assessments) of concordance between systematic reviews addressing similar questions, and between practice guideline recommendations addressing similar health topics. Eligible studies may report on the prevalence of concordance, or may evaluate reasons for discordance. Studies may address one or multiple medical conditions.

We will also include concept papers (i.e. reporting methodological frameworks) proposing methods or approaches to perform concordance assessments or evaluations of reasons for discordance.

We will exclude individual systematic reviews and individual clinical practice guidelines; we will limit eligibility to evaluations of concordance within both study designs. Reports of systematic reviews or practice guidelines that discuss concordance of findings or recommendations with others’ only in their discussion or interpretation will be ineligible. We will also exclude assessments of concordance involving different study designs (e.g. systematic review of observational studies versus systematic review of randomized trials), grey literature, and other study designs (including editorials, commentaries, correspondences, letters to editors, news articles).

### Search strategy

We will conduct an electronic database search using OVID MEDLINE, Embase and CINAHL from inception to June 2022 using medical subject heading (MeSH) terms and keywords related to the following two concepts: (1) systematic reviews (including meta-analyses); guidelines (including guidance and recommendations); and (2) concordance (including discordance, discrepancy, agreement, convergence, divergence, disagreement and consistency).

We will supplement our electronic database search by reviewing bibliographies of identified articles, and consulting experts for additional potentially eligible studies not identified by the electronic searches.

### Study selection

Paired reviewers will independently conduct title and abstract and full text screening, with conflicts resolved by discussion, and, if needed, with input from a third reviewer. Screening will be conducted using Covidence. Reviewers will complete a pilot exercise for training and calibration before each stage of screening.

### Data extraction

Paired reviewers will use standardized forms to independently extract data from eligible studies, following an initial pilot exercise. They will resolve discrepancies by discussion, and, if needed, with input from a third reviewer.

We will collect the following data from each study:

- Bibliometric information: first author, year of publication, journal.
- General characteristics of the study: type (empirical study vs. concept paper), stated objective, number of systematic reviews or guidelines assessed, number of outcomes or recommendations assessed.
- Characteristics of assessed reviews or guidelines; field (clinical, public health, health systems), type (update, living, rapid; for guidelines, de novo or adapted), disease of interest, PICOs addressed.
- Steps of the concordance assessment process studied: method of identification of referent systematic review (and associated PICOs or outcomes) or guideline (and associated recommendations), method of identification of related systematic reviews or guidelines, matching of PICOs or recommendations, assessment of concordance, evaluation of reasons for discordance.
- Results of quantitative analyses (e.g., proportions of concordance, reasons for discordance).
- Results of any qualitative analyses.

### Data analysis

We will synthesize data quantitatively where possible; where not possible, we will use a narrative approach. For continuous variables, we will use means and standard deviations for normally distributed variables, and medians and interquartile ranges for non-normally distributed variables. For categorical variables, we will use frequencies and percentages.

We ultimately plan to use our findings to ultimately develop an evidence-based framework for assessment of concordance between systematic reviews, and between clinical guidelines.

## Data Availability

All data produced in the present work are contained in the manuscript.

## Data availability

All data produced in the present work are contained in the manuscript.

